# Development and Psychometric Testing of the Maternity Empowerment (MPower) instrument

**DOI:** 10.64898/2026.05.14.26353220

**Authors:** Kate Buchanan, Alisha Kaumanns, Lukman Thalib, Patricia Leahy-Warren, Marianne Nieuwenhuijze

**Affiliations:** School of Nursing and Midwifery, Edith Cowan University, Perth, Western Australia, Australia; Research Centre for Midwifery Science, Zuyd University of Applied Sciences, Maastricht, The Netherlands; Department of Biomedical Engineering, Yildiz Technical University, Istanbul, Turkiye; School of Nursing and Midwifery, University College Cork, Cork, Ireland; Care and Public Health Research Institute (CAPHRI), Maastricht University, Maastricht, The Netherlands

**Keywords:** Midwifery, Perinatal, empowerment, midwifery models of care, validation, instrument

## Abstract

**Introduction:** Perinatal Empowerment is widely referenced in maternity care research, yet its use often lacks clear conceptual definitions and validated measures. Existing instruments do not capture the multidimensional nature of perinatal empowerment, including both external dimensions (e.g., gender equity, resource access), and internal dimensions (e.g., confidence, agency and informed decision making). This gap has limited the ability to rigorously evaluate how healthcare experiences shape empowerment during pregnancy, birth, and the postpartum period.

**Aim:** To develop a valid and reliable instrument that measures dimensions of perinatal empowerment, both external and internal.

**Methods:** Instrument development followed the seven-step MEASURE framework. Initial item generation was guided by a concept analysis, a scoping review of existing instruments, and feedback from international midwifery experts. A preliminary 51-item instrument underwent expert content validity review, resulting in 48 items, which were then pilot-tested with six pregnant and postnatal women. A large-scale validation study was conducted via an international online survey (N=155). Psychometric testing included exploratory factor analysis (EFA), reliability assessment using Cronbach’s α, known-groups validity testing, and regression analyses adjusting for potential confounders.

**Results:** EFA supported two overarching dimensions—external and internal empowerment—with six factors across 30 final items (18 external, 12 internal). Sampling adequacy was high, and item loadings exceeded recommended thresholds. Internal consistency was strong for both dimensions (α=0.88 external; α=0.87 internal). Women receiving midwifery continuity of care reported significantly higher empowerment scores across total, external, and internal dimensions compared with other care models (p<.001). Differences between primiparous and multiparous women were not statistically significant.

**Conclusion:** The MPower instrument represents a conceptually grounded, psychometrically robust measure of multidimensional perinatal empowerment in high-income settings. Further validation in more diverse populations is needed to refine the instrument and expand its applicability across clinical and research contexts.

## Introduction

The perinatal period encompassing pregnancy, birth and postpartum period represent a critical life stage characterised by profound physical, emotional, and psychological changes (1), where empowerment seems to be one factor that enables women to navigate these transformations positively to their new life-situation (2, 3). Across maternity research and practice, empowerment is frequently proposed as a desirable outcome of respectful care (4), yet is often used as a broad, unexamined label that lacks a consistent definition and is rarely supported by robust measurement (5). This widespread but uncritical use obscures the fact that empowerment, and existing claims about its effects, are not grounded in validated evidence. Consequently, empirical evidence capturing the full scope of perinatal empowerment remains incomplete, highlighting a clear need for a reliable and conceptually sound instrument to assess this construct.

Empowerment is widely regarded as a multidimensional construct (6, 7), resulting in diverse definitions across the fields of demography, economics, and public health (8, 9). The absence of a unified definition poses a challenge in conceptualising women’s empowerment within the perinatal context (6). Conceptual ambiguity has direct influence for its measure (10). Consequently, there can be considerable variability in the association between reproductive empowerment and health outcomes. Kabeer’s 1999 (11) foundational work on women’s empowerment, defines it as the ability to make strategic and meaningful life choices across socio-cultural, economic, and psychological dimensions. Expanding on this, Nieuwenhuijze and Leahy-Warren (6) conducted a concept analysis that identified two categories of attributes specific to perinatal empowerment (Box 1). External attributes include gender equity, access to and control over resources, and support for women’s autonomy in decision-making during the perinatal period. Internal attributes encompass women’s belief in their own abilities, self-control, and perceived capacity to have control over others and situations (6).

#### Box 1. Defining attributes of empowerment

**External attributes**

- Gender equality
- Access and control of resources
- Facilitation of women’s choice and decisions

**Internal attributes**

- Women’s belief in own abilities
- Control over situation, self and others

Building on this concept analysis, Leahy-Warren and Nieuwenhuijze (12) undertook a scoping review of instruments designed to measure perinatal empowerment. The review retrieved 19 instruments used to assess perinatal empowerment and related concepts, however, only six of these had undergone formal validation. The findings identified substantial variability across instruments, with many failing to measure both internal and external attributes of empowerment; several omitted all internal attributes (KAS-R, MADM, Repro-Q/MEQ) while others omitted all external attributes (Empowerment Score, Levenson Scales), and key elements such as access to resources and social support were frequently excluded. Additionally, many instruments focused narrowly on a single perinatal stage or dimension of empowerment, resulting in no tool capturing all defining attributes identified in the concept analysis (6). This current study therefore builds on this foundational work to progress the development of a preliminary instrument, addressing a long-standing gap in the field and moving towards a more rigorous, evidence-based understanding of empowerment. To address the absence of a comprehensive instrument that encompasses all relevant dimensions and attributes of empowerment, this current study delineates the development of such an instrument.

**Aim:** Our aim was to develop a valid and reliable instrument that measures dimensions of perinatal empowerment, both external and internal.

## Methods

### Ethical statement

Approval to conduct this study was obtained from a Human Research Ethics Committee at Edith Cowan University, (HREC 2024-0560). All participants provided free and written informed consent and remained anonymous and unidentifiable throughout the research process. The study adhered to the ethical principles outlined in the Declaration of Helsinki and followed best-practice publication standards consistent with the Committee on Publication Ethics (COPE), including expectations regarding authorship, originality, data integrity, and responsible reporting (STROBE).

The research design followed the MEASURE Approach for instrument development (13), a framework comprising seven empirically supported steps. These steps include: 1. **M**ake clear purpose and rational; 2. **E**stablish empirical framework; 3. **A**rticulate theoretical blueprint; 4. **S**ynthesize content and instrument development; 5. **U**se expert reviewers; 6. **R**ecruit participants; **7. E**valuate validity and reliability. The following section describes each step in detail.

#### Step 1. Purpose and rational for instrument development

A theoretically grounded, valid, and reliable instrument that captures the full range of empowerment dimensions experienced by women during the perinatal period is needed to generate evidence that will guide the design of coherent, theory-informed, and empirically supported interventions to strengthen perinatal empowerment. A scoping review of empowerment instruments by Leahy-Warren and Nieuwenhuijze, (2023) synthesised existing literature on perinatal empowerment instruments concluded that no available instrument captured the dimensions of empowerment as both external and internal attributes of women’s empowerment. The review emphasised the need for a comprehensive instrument that aligns fully with the multidimensional definition of empowerment.

#### Step 2 and 3. Empirical framework and theoretical blueprint

A theoretical blueprint is essential to ensure that an instrument comprehensively covers all relevant attributes of the construct. The concept analysis by Nieuwenhuijze & Leahy-Warren, (2019) outlined in the background section (see Box 1) identified two dimensions of empowerment: external and internal. Together, these dimensions provide the empirical framework and theoretical foundation required for instrument development and characterise the attributes of empowerment in the perinatal period (6). External attributes encompass gender equality, access to and control over resources, and the facilitation of women’s autonomy and decision-making. Internal attributes include women’s belief in their own abilities, self-control, and perceived capacity to influence situations and others. These attributes formed the empirical foundation from which the dimensions and preliminary items were generated, providing a coherent, literature-based conceptual structure to guide subsequent instrument refinement. The theoretical framework ensured that the instrument fully reflects the multidimensional nature of empowerment during the perinatal period.

#### Step 4. Synthesise content and instrument development

The scope and depth of the content was ensured through a comprehensive review of the literature. An initial pool of items extracted from existing instruments found in the scoping review were crafted under the dimensions (external and internal) and attributes in the framework based on the concept analysis of perinatal empowerment (6). An initial draft was discussed with a group of 11 international stakeholders from midwifery and Public Health Nursing at a workshop at the Normal Birth Conference 2023. Participants provided feedback on the relevance and wording of possible items and resulted in a first draft of the instrument.

Over the course of a year, Buchanan, Nieuwenhuijze, Leahy-Warren and, bringing together diverse expertise in midwifery research and clinical practice further developed the instrument items through a rigorous, iterative process involving multiple rounds of review and refinement. Each iteration involved a thorough examination of the items, focusing on their clarity, relevance, and alignment with empowerment. This iteration, resulting in a 51-item instrument, was ready for examining content validity using expert review (Step 5).

Likert scaling was chosen for the instrument, as its one of the most used scaling formats in the social sciences and appropriate for measuring attitudinal constructs (e.g., personality, beliefs, values, or emotions) (14). The items were presented as declarative statements, designed for varying amounts of agreement with the statement, for example, 1= strongly disagree, 2= disagree, 3= neutral, 4= agree, 5= strongly agree.

#### Step 5. Use expert reviewers

An assessment on the instrument’s content validity was conducted in June 2024 by nine expert reviewers. They were purposely selected international midwifery leaders, including tertiary hospital midwifery managers, national college chief executives, midwifery academics, and midwives with a private practice. Expert reviewers were asked to complete an online survey and rate on a Likert scale the extent to which each of the questions demonstrated the characteristics of empowerment, relevance to measuring empowerment, and clarity of the item wording. Responses were recorded on a seven-point Likert scale ranging from Strongly Disagree (1) to Strongly Agree (7). Open-response options were available for comments for each item. The total sum score for each item was calculated (range 9-63). Any item with a lower sum score of 45 was reviewed and wording was adjusted as appropriate, cognisant of comments given. Three items were removed due to consistently low scores across all three areas, resulting in a 48-item instrument.

#### Step 6. Recruit participants Pilot testing

The instrument required pilot testing with end users to determine comprehensibility of the instrument. Subsequently, this 48-item instrument was pilot tested in October 2024 with six multiparous women: two who were pregnant and four who were postnatal, including one participant for whom English was a second language. A convenience sampling approach was used, reaching out to consumer groups and collegiate networks, to distribute an advertisement via email including an information letter and an online consent form. Interested participants who consented were scheduled for an online meeting at a mutually convenient time. For a pilot study using interviews, it is commonly agreed that a sample size of 6–10 participants is sufficient (15). Variation in ethnicity, location, education, parity, age, and model of care were achieved. Six participants completed the instrument online with a researcher (first author), giving verbal feedback on each item as a “Think-out-loud” exercise (16). The interviews were recorded and transcribed verbatim.

The transcripts were reviewed for any participant confusion, leading to amendments in the wording of three items. No items were removed.

This completed the content validity process of the instrument, during which issues related to item clarity were identified and addressed, thereby ensuring the overall comprehensibility of the instrument.

#### Step 7. Evaluate validity and reliability

##### Validation study

Psychometric evaluation of the instrument was conducted through an international online survey. Women were recruited via electronic advertisements disseminated through social media, consumer groups, and professional networks. Participants accessed the survey through a link to the information sheet and consent form, followed by eligibility screening and signing the electronic consent form. Eligible participants were women aged ≥18 years, proficient in English, and either pregnant or within one year postpartum. Data collected were hosted on Qualtrics™, which enabled secure administration and direct export of responses for analysis. Data were collected from December 2024 to February 2025.

Sample size was determined a priori using the subject-to-item ratio of 3–5 participants per item (17). With 48 items, the minimum requirement was 144 participants; to allow for attrition, a target of 180 participants was set.

The survey included demographic questions on birth-related characteristics (e.g., age, marital status, education, parity, model of care, labour onset, and birth mode), followed by the 48-item instrument. Items were rated on a five-point Likert scale, with 10 items reverse-scored to assess response consistency. Total scores ranged from 48 to 240, with higher scores reflecting greater empowerment.

##### Data Analysis

Psychometric evaluation involved testing for validity (the instrument is measuring what it is intended to measure) and reliability (consistency of scores) evidence of the instrument and its subscales (18). Data were analysed using several descriptive, psychometric, and inferential statistical tests.

Demographic characteristics were analysed using descriptive statistics (frequencies and percentages) (19).

Total scores on the instrument were computed by summing up all item responses. For both the external and internal dimensions, scores were calculated by summing up the corresponding item responses. Missing data within the items were addressed using multiple imputation. Prior to the imputation, a missing data analysis was conducted to assess any patterns in the missing values. Results indicated that the data were missing at random, supporting the use of multiple imputation. Exploratory factor analyses, construct validity testing and calculating Cronbach coefficient alpha were conducted in SPSS Statistics 30.

##### Exploratory factor analysis (EFA)

Exploratory factor analysis (EFA) was conducted to examine the factor structure of the 48-item instrument. EFA identified factors within both the external and internal dimensions by detecting correlations among groups of items, each representing an aspect of the overall construct being measured. The analysis included evaluation of the inter-item correlation matrix, factor extraction, factor rotation, factor retention, and the naming of the rotated factors.

Items with factor loadings ≥ 0.40 on at least one factor were retained. Items demonstrating cross-loadings on two or more factors, with a difference in loadings of ≤ 0.30, were removed following team discussion.

##### Reliability analysis

Reliability was evaluated using standard psychometric methods. Internal consistency was assessed by calculating Cronbach’s alpha for both the external and internal dimensions after initial pilot testing. Items with low corrected item–total correlations or evidence of cross-loading were reviewed and either modified or removed as necessary to improve overall reliability.

##### Construct validity

The construct validity of the instrument was assessed in terms of known group validity. We formulated two hypotheses, one linking to the external dimension of empowerment, and one linking to the internal dimension of empowerment. Related to external dimensions we hypothesized that women receiving Midwifery Continuity of Care (MCoC), during pregnancy and birth, have higher total, external and internal scores on perinatal empowerment than women who received care in other care models (Hypothesis 1) (20–23). Women’s sense of empowerment is positively linked to their involvement in decision-making and their satisfaction with the care received during pregnancy and childbirth (24–27). Women-centered care models, like MCoC, support women’s choice and are likely positively associated with a higher sense of empowerment (2, (28). Furthermore, MCoC is positively associated with women’s sense of empowerment by providing continuous, woman-centered support, which enhances women’s confidence, promotes informed decision-making, and reduces anxiety during pregnancy and childbirth (20–23).

Related to internal dimensions, we hypothesized that women who are currently pregnant with their first child or have given birth once (named primiparous women) have a lower total empowerment score, and a lower score on the external and internal empowerment dimensions than women who have given birth to more than one child (named multiparous women) (Hypothesis 2). We made this assumption based on studies that measured self-efficacy and confidence (29–31) as no studies seem to exist that measure the full concept of perinatal empowerment (14).

These analyses were done with the data of the 30 items that remained in the instrument after the factor analyses (18 items of the External dimension and 12 items of Internal dimension). A Kruskall-Wallis test examined differences in perinatal empowerment scores based on model of care (Hypothesis 1), as data were not normally distributed, and this involved comparisons across more than two groups. Median scores and interquartile range are reported in the results section. We examined differences in perinatal empowerment scores based on parity (Hypothesis 2) using Mann-Whitney U test, as data were not normally distributed.

To extend the analyses and rule out any possible confounding, we subsequently examined demographic and clinical variables, that could affect women’s sense of empowerment. Based on previous research, the following variables were identified as potential co-variates in affecting sense of empowerment: maternal age, parity, relationship status, educational level, birth mode and model of care (29, 32–36). These variables were included in a logistic regression analysis to explore which variables contribute to women’s empowerment in the perinatal period. A p-value <0.05 was considered statistically significant.

#### Findings

##### Participant characteristics

A total of 168 women initiated the survey. Of these, 155 women completed the study and were included in the final analysis. Thirteen participants were excluded due to incomplete data, including 10 without survey data, 2 with more than two missing items on the instrument, and 1 without data on main healthcare provider during the last pregnancy. The majority of participants resided in Australia (n=113), while all others, except one lived in high-income countries. Most participants were also born in high-income countries, with the exception of three. The most common age group was 31-35 years (n=64) and the majority had completed university (n=131). Approximately 45% of participants (n=69) were primiparous women, and 42.6% (n=66) reported receiving continuous midwifery care during their last pregnancy, birth and postpartum (see Table 1.).

**Table 1:**
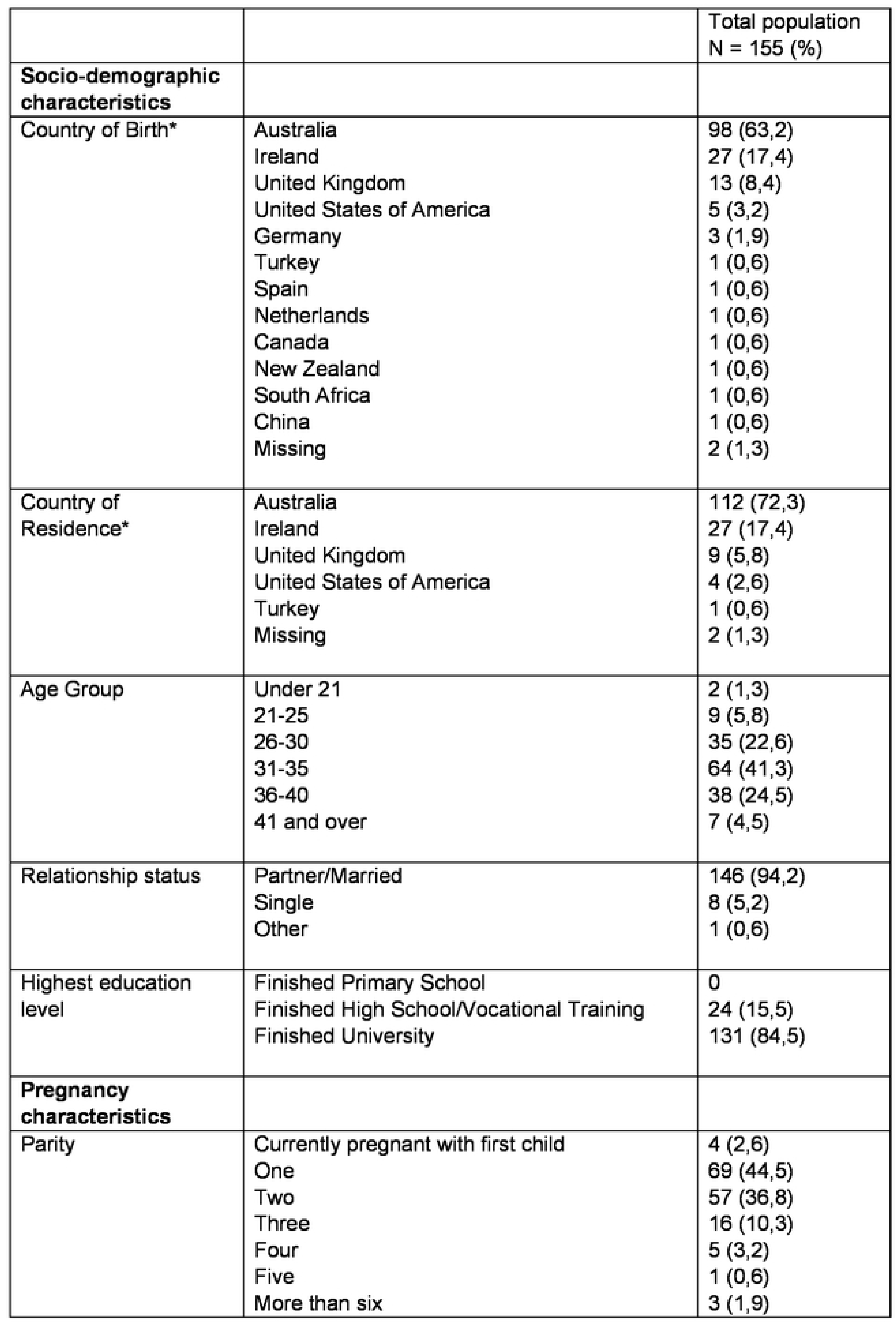
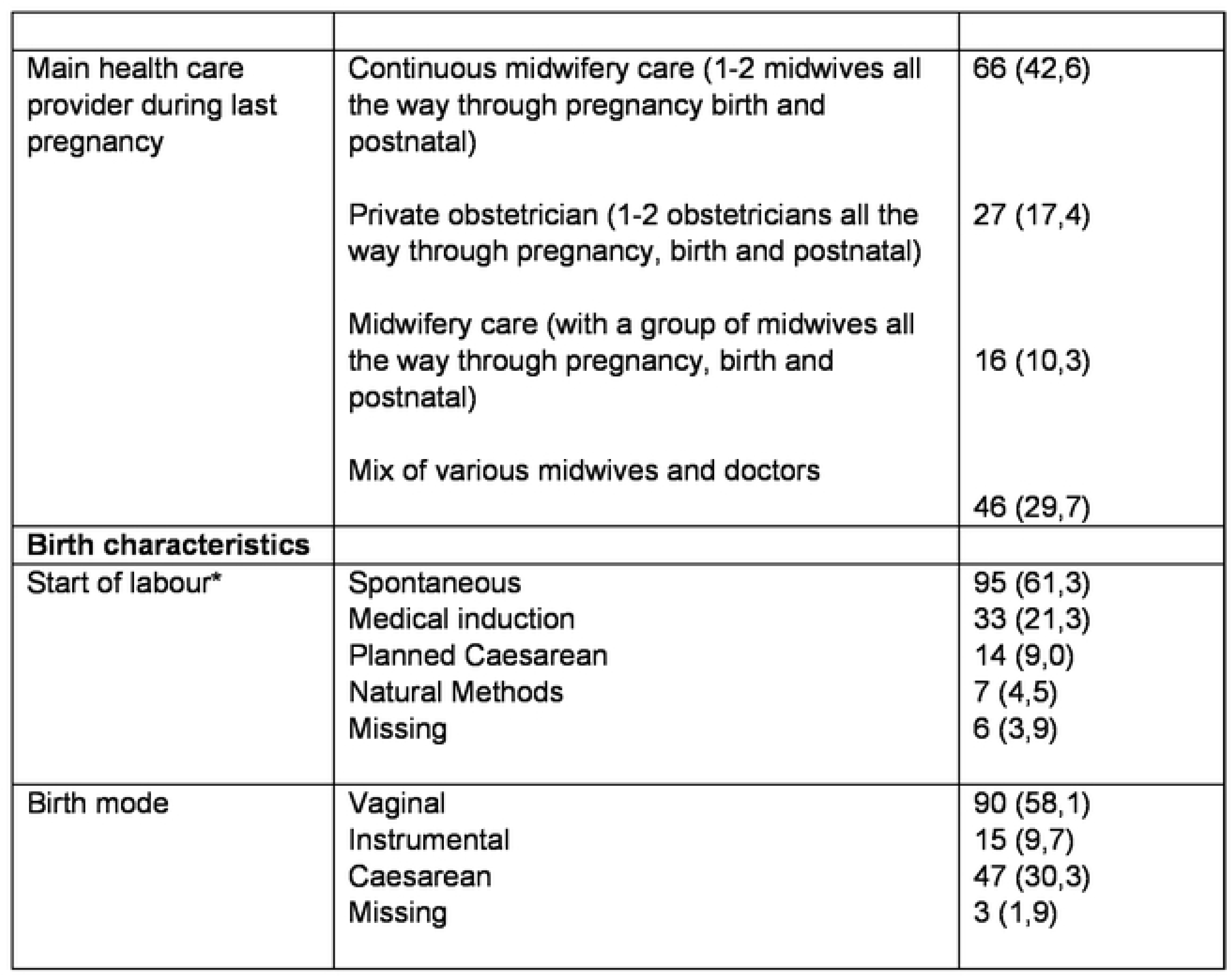
Baseline participant characteristics. * Some demographic data are incomplete due to missing responses

##### Factor analysis

An Exploratory Factor Analysis (EFA) was conducted to examine the underlying structure of the instrument. The analysis was performed separately for the two hypothesized dimensions: external empowerment and internal empowerment. Sampling adequacy was verified using the Kaiser–Meyer–Olkin (KMO) measure and Bartlett’s test of sphericity. The KMO values for the external and internal dimensions were 0.899 and 0.885, respectively, indicating sampling adequacy. Bartlett’s tests were significant (p < 0.001), confirming the factorability of the data.

External Empowerment Dimension

For the external dimension, a three-factor solution emerged, explaining 63% of the variance. Factor loadings ranged from **0.54 to 0.89** (Table 2). The identified factors were interpreted as collaborative care (factor E,1), societal enablement (factor E.2) and reproductive equality (factor E.3).

**Table 2:** Factor Loadings for External Empowerment Dimension.

| External empowerment item | Factor 1 | Factor 2 | Factor 3 | Communality |
| --- | --- | --- | --- | --- |
| **Collaborative care** |  |  |  |  |
| *<u>My main health care provider:</u>* |  |  |  |  |
| ensured my preferences were a priority | 0.87 | - | - | 0.80 |
| kept me informed about what was happening during care | 0.86 | - | - | 0.75 |
| made me comfortable declining care that was offered | 0.86 | - | - | 0.74 |
| took my concerns seriously | 0.86 | - | - | 0.73 |
| gave me enough time to discuss my care | 0.85 | - | - | 0.73 |
| respected my choice once I made a decision | 0.84 | - | - | 0.70 |
| made me feel inferior (like I were less important than they were) | 0.78 | - | - | 0.63 |
| ensured that I could get all my questions answered | 0.77 | - | - | 0.64 |
| invited me to express my own cultures beliefs, customs and habits | 0.75 | - | - | 0.58 |
| informed me that there were different options for my maternity care | 0.69 | - | - | 0.51 |
| always welcomed a significant other to be with me | 0.55 | - | - | 0.30 |
| **Societal enablement** |  |  |  |  |
| In my community, women can make their own choices about whether they become pregnant or not | - | 0.75 | - | 0.57 |
| In my country or state, laws and regulations are equal for all sexes | - | 0.71 | - | 0.60 |
| In my community, women have easy access to resources, such as transport, money and health services | - | 0.69 | - | 0.49 |
| In my community, women experience discrimination because of their pregnancy | - | 0.61 | - | 0.39 |
| **Reproductive agency** |  |  |  |  |
| The health system in my country or state, ensured I knew what my rights were | - | - | 0.89 | 0.82 |
| The health system in my country or state, provided resources (e.g. staff, leaflets) that helped to make my own decision | - | - | 0.88 | 0.83 |
| In my country or state, women have full human rights over their body and decisions about their body | - | - | 0.54 | 0.57 |
| Cronbach's $\alpha$ = 0.88 | | | | |

Internal Empowerment Dimension

EFA using varimax rotation revealed a clear three-factor solution, explaining 65% of the total variance. Items loaded strongly on their respective factors, with loadings ranging from 0.65 to 0.84 (Table 2). Factor I.1 reflected sense of agency, factor I.2 self-belief, while Factor I.3 represented self-reliance.

**Table 3.** Factor Loadings for Internal Empowerment Dimension.

| Internal empowerment Item | Factor 1 | Factor 2 | Factor 3 | Communality |
| --- | --- | --- | --- | --- |
| **<u>Sense of Agency</u>** |  |  |  |  |
| I had an equal partnership with my care provider | 0.84 | - | - | 0.79 |
| I felt confident (trusted) in my care provider because they knew me | 0.82 | - | - | 0.67 |
| If I had wanted to, I could change the care I received | 0.79 | - | - | 0.69 |
| I was in control over decisions made about my care | 0.78 | - | - | 0.78 |
| I understood what was happening to me | 0.76 | - | - | 0.70 |
| I had no control over who touched my body | 0.67 | - | - | 0.50 |
| **<u>Self-belief</u>** |  |  |  |  |
| I am confident I could help to solve problems associated with my pregnancy, birth and postpartum | - | 0.76 | - | 0.65 |
| I am the person who is responsible for taking care of my health | - | 0.76 | - | 0.59 |
| If a situation became difficult, I could manage it | - | 0.68 | - | 0.71 |
| Pregnancy, birth and postpartum experiences made me stronger | - | 0.65 | - | 0.48 |
| **<u>Self-reliance</u>** |  |  |  |  |
| During my care I wished someone else would make decisions on my behalf | - | - | 0.81 | 0.67 |
| I was comfortable receiving care from unknown care providers who didn't introduce themselves | - | - | 0.75 | 0.58 |

Cronbach's $\alpha$ = 0.87

Based on the EFA we removed 11 items from the external dimension and seven items from the internal dimension, leaving an instrument with 30 items (18 external and 12 internal).

##### Reliability Analysis

The overall reliability coefficient measured via Cronbach’s α for external empowerment dimension was 0.88, while the corresponding α for internal empowerment dimension was 0,87. indicating strong internal consistency across the instrument.

In summary, EFA supported a multidimensional structure of the instrument, comprising two overarching dimensions (external and internal empowerment) and six factors. Both dimensions demonstrated satisfactory internal consistency, with Cronbach’s α values exceeding the recommended threshold of .70.

##### Construct validity

Analysis for hypothesis one (H1) revealed a statistically significant difference in empowerment scores between the different models of care (*p* <.001) (Hypothesis 2) (Table 4). Median total perinatal empowerment scores were highest among women receiving MCoC (median=132), compared to a group of midwives all the way thought pregnancy, birth and postnatal (median=101) and other models of care (median=118). On the external dimension, women receiving MCoC also scored statistically significant (*p<.*001) higher (median=76) than women in group midwifery care (median=61) and other models of care (median=69). Similarly, on the internal dimension, women receiving MCoC scored statistically significant (*p<.001)* higher (median=56) than women in group midwifery care (median=41) and women in other models of care (median=46). These findings suggests that women in MCoC experience a higher sense of empowerment than those in other models of care.

**Tabel 4.** Empowerment scores for women receiving MCoC, group midwifery care and other models of care.

|  | n | Total score |  | External subscale |  | Internal subscale |  |
| --- | --- | --- | --- | --- | --- | --- | --- |
|  |  | Median (interquartile range) | p-value | Median (interquartile range) | p-value | Median (interquartile range) | p-value |
| MCoC<sup>a</sup> | 66 | 132 (124, 138) |  | 76 (70, 81) |  | 56 (53, 59) |  |
| Group midwifery Care<sup>a</sup> | 16 | 101 (87,120) |  | 61 (55, 69) |  | 41 (36, 49) |  |
| Other models of care<sup>b</sup> | 73 | 118 (104, 131) |  | 69 (62, 79) | <.001 | 46 (40, 53) |  |
|  |  |  | <.001 |  |  |  | <.001 |

<sup>a</sup> Continuous midwifery care (1-2 midwives)
<sup>b</sup> Care given by a group of midwives
<sup>c</sup> Care given by (private) obstetrician or a mix of various doctors and midwives

Analysis for hypothesis two (H2) (Table 5) revealed that multiparous women had a higher median total perinatal empowerment score (median=126), compared to primiparous women (median=120). However, this difference was not statistically significant (*p*=.25). There was also no statistically significant difference in scores either on the external dimension between multiparous women (median=73) and primiparous women (median=71; *p*=.60), nor on the internal dimension where multiparous women scored higher (median=53) than primiparous women (median=49; *p*=.12).

**Table 5.** Empowerment scores for primiparous and multiparous women.

|  | n | Total score |  | External subscale |  | Internal subscale |  |
| --- | --- | --- | --- | --- | --- | --- | --- |
|  |  | Median (interquartile range) | p-value | Median (interquartile range) | p-value | Median (interquartile range) | p-value |
| Primipara | 73 | 120 (106, 132) |  | 71 (62,80) |  | 49 (41,56) |  |
| Multipara | 82 | 126 (112, 134) |  | 73 (65,79) |  | 53 (45,58) |  |
|  |  |  | 0.25 |  | 0.60 |  | 0.12 |

After adjusting for potential confounders (maternal age, parity, relationship status, educational level and birth mode), linear regression analysis showed that women in other models of care (β=-13.73, 95% CI [-20.06, -7.39], *p*<.001) and in group midwifery care (β=-26.97, 95% CI [-37.08, -16.86], *p* <.001) have statistically significant lower total perinatal empowerment scores compared to women receiving MCoC. On the external dimensions women in other models of care and women in group midwifery care also had statistically significant lower scores than women in continuous midwifery care (Table 6). There was no statistically significant difference in total empowerment scores, external dimensions and internal dimensions between primiparous and multiparous women after adjustment for co-variates (Table 6).

**Table 6.** Linear Regression.

| Predictor | Total score |  |  | External subscale |  |  | Internal subscale |  |  |
| --- | --- | --- | --- | --- | --- | --- | --- | --- | --- |
|  | B | 95% CI | p | B | 95% CI | p | B | 95% CI | p |
| Parity (multiparous vs. primiparous) | 0.15 | (-5.96, 6.26) | 0.96 | 0.03 | (-4.16, 4.21) | 0.99 | 0.13 | (-2.47, 2.72) | 0.92 |
| Maternal age |  |  |  |  |  |  |  |  |  |
| Age < 21 year | -3.93 | (-30.56, 22.69) | 0.77 | -2.93 | (-21.17, 15.32) | 0.75 | -1.01 | (-12.32, 10.31) | 0.86 |
| Age 21-25 year | 2.21 | (-10.84, 15.26) | 0.74 | 1.10 | (-7.85, 10.05) | 0.81 | 1.11 | (-4.44, 6.65) | 0.69 |
| Age 26-30 year | -3.41 | (-11.04, 4.23) | 0.38 | -0.79 | (-6.02, 4.45) | 0.77 | -2.62 | (-5.87, 0.62) | 0.11 |
| Age 31-35 year* |  |  |  |  |  |  |  |  |  |
| Age 36-40 year | 0.24 | (-7.06, 7.54) | 0.95 | 0.80 | (-4.20, 5.81) | 0.75 | -0.56 | (-3.67, 2.54) | 0.72 |
| Age > 41 year | -2.14 | (-16.34, 12.07) | 0.77 | -3.09 | (-12.83, 6.64) | 0.53 | 0.96 | (-5.08, 7.00) | 0.75 |
| Relationship status |  |  |  |  |  |  |  |  |  |
| Partner/married | 8.66 | (-4.32, 21.65) | 0.19 | 3.63 | (-5.27, 12.53) | 0.42 | 5.03 | (-0.49, 10.55) | 0.07 |
| Single/other | -12.19 | (-49.49, 25.11) | 0.52 | -13.35 | (-38.91, 12.22) | 0.30 | 1.16 | (-14.69, 17.00) | 0.89 |
| Educational level (high vs middle) | -0.71 | (-9.00, 7.58) | 0.87 | -1.41 | (-7.09, 4.27) | 0.62 | 0.70 | (-2.82, 4.23) | 0.69 |
| Birth mode |  |  |  |  |  |  |  |  |  |
| Spontaneous vaginal* |  |  |  |  |  |  |  |  |  |
| Instrumental birth | -6.94 | (-17.28, 3.39) | 0.19 | -2.08 | (-9.16, 5.01) | 0.56 | -4.87 | (-9.26, -0.48) | **0.03** |
| Cesarean section | -5.62 | (-12.57, 1.33) | 0.11 | -1.34 | (-6.10, 3.42) | 0.58 | -4.28 | (-7.23, -1.32) | **<0.01** |
| Model of care |  |  |  |  |  |  |  |  |  |
| Other model of care (vs. continuous midwifery care) | -13.73 | (-20.06, -7.39) | **<.001** | -6.33 | (-10.67, -1.99) | **<0.01** | -7.40 | (-10.09, -4.71) | **<.001** |
| Group midwifery care ^ (vs. Continuous midwifery care) | -26.97 | (-37.08, -16.86) | **<.001** | -14.18 | (-21.11, -7.25) | **<.001** | -12.79 | (-17.09, -8.50) | **<.001** |

\* reference group
^ defined as a group of midwives all the way through pregnancy, birth and postnatal
Statistically significant results ( $p \leq 0.05$ ) are shown in bold

##### Instrument name: MPower

Following validation, the instrument was named the Maternity empowerment (MPower) instrument comprising two dimensions containing a total of 30 items. The MPower (Box 2) consists of 30 items organised into two dimensions. The external dimensions include 18 statements addressing gender equality, access to and control of resources, and the facilitation of women’s choice and control. The internal dimension comprises 12 statements related to women’s beliefs in their own abilities, control over situations, and influence over themselves and others. MPower offers a structured assessment of perinatal empowerment across both external and internal dimensions.

###### Box 2. MPower - Maternity empowerment instrument

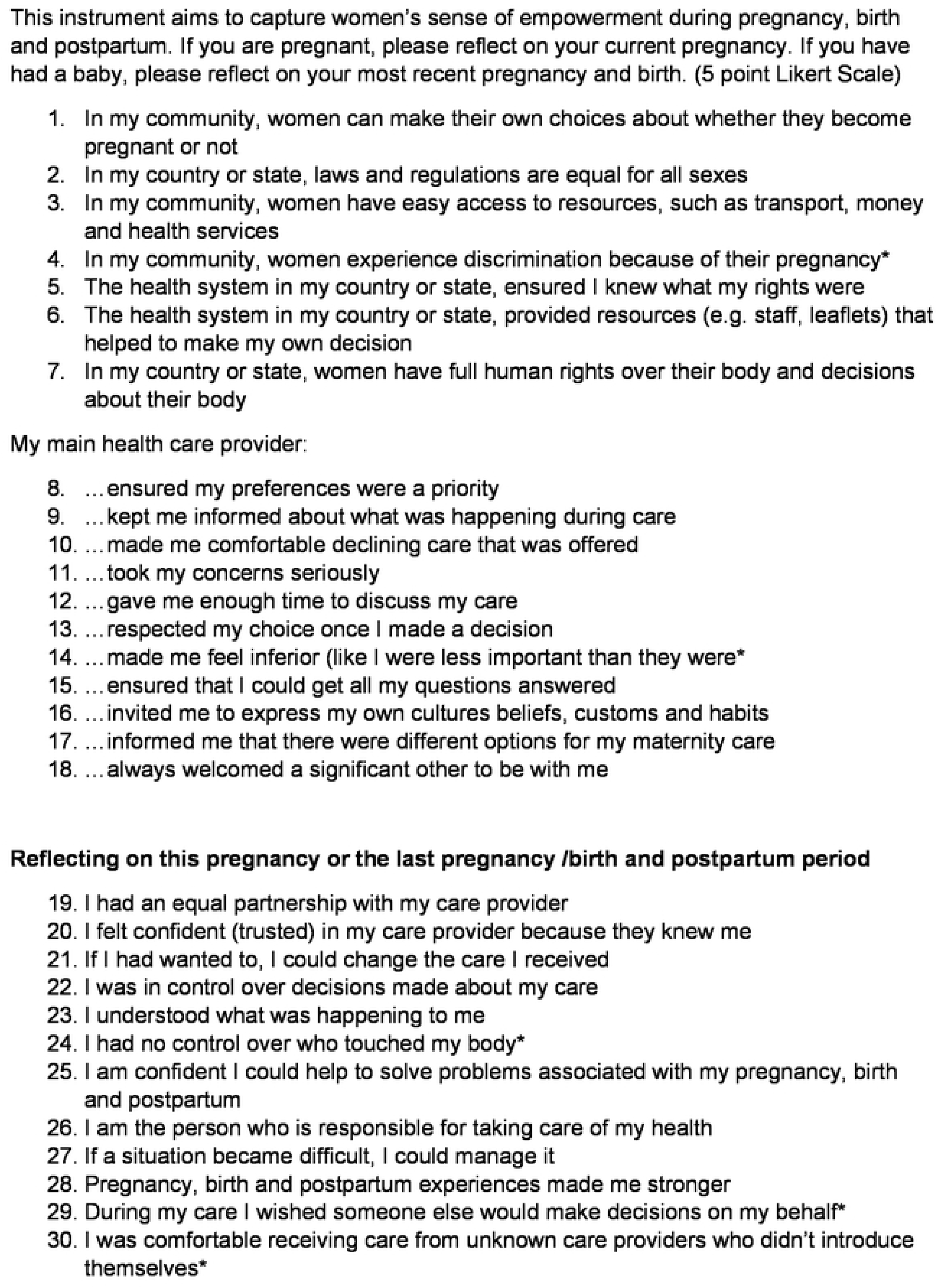

## Discussion

Although empowerment is widely invoked within the maternity care literature, it is often treated as a rhetorical device rather than an evidence-based construct, with its significance emphasised in the absence of rigorous conceptualisation or validated measurement. This persistent gap has resulted in inconsistent and unsubstantiated use of the term, limiting its scientific and practical utility. Our programme of work addresses this deficit by systematically identifying the external and internal dimensions that underpin perinatal empowerment and used these as the foundation for the development of a preliminary instrument. While this represents meaningful progress toward a robust measure, the instrument remains at an early stage of development and requires further testing, validation, refinement, and likely item reduction before it can be considered complete. This study therefore contributes to an essential and ongoing process of bringing conceptual clarity and empirical rigour to the assessment of empowerment in the perinatal context.

The MPower instrument comprises 30 items organised into two dimensions of empowerment: external and internal. It can be administered either as a single instrument or focusing on one dimension depending on purpose. Initial psychometric analyses suggest the instrument captures variance across both dimensions with promising properties; however, these findings are preliminary. Further research with larger, more diverse samples, formal validation across settings, iterative refinement, and potential item reduction is needed before strong conclusions can be drawn or routine use recommended.

Using our instrument, our findings identified that women receiving continuous midwifery care reported higher empowerment scores compared with those receiving other models of care, even after adjusting for covariates. These results align with broader evidence linking midwifery continuity of care with enhanced empowerment. Oglak and colleagues (2026)(37) similarly demonstrated that continuity of midwifery care fosters empowerment, measured using the Pregnancy Empowerment Scale (PES): a 13-item, four-point Likert instrument developed by Klima and colleagues (2015)(38), although only assessing four subdomains of empowerment: Provider Attachment, Competent Decision-Making, Peer Attachment, and Voice. Higher PES scores reflect greater empowerment, and women in the intervention group reported significantly higher empowerment and health literacy, with the two positively correlated (r = .748).

Given the aims and design of this study, no conclusions can be drawn regarding associations between empowerment and other factors. Exploring such relationships was beyond the scope of this work. Further research, with larger and more diverse populations, is required to understand which conditions, circumstances, or interventions may be correlated with empowerment, and different study designs will be necessary to establish any causal pathways. Similarly, additional research is needed to examine how empowerment may influence outcomes during the perinatal period and beyond. While this instrument contributes one component to advancing inquiry in this area, much remains to be understood about how women can be better supported to feel empowered throughout the perinatal journey. For healthcare providers, developing a clearer evidence base regarding the relationship between care practices and women’s empowerment will be essential to informing care that promotes positive and enduring outcomes.

### Strengths and Limitations

This study’s strengths include the rigorous development process of the content for the instrument, which involved a scoping review, empirical framework establishment, content synthesis, expert review, pilot testing, and psychometric evaluation. The MPower scale is the first instrument to include external and internal dimensions of perinatal empowerment which provides a comprehensive understanding of empowerment during the perinatal period. The input from the end users via the pilot study strengthens the instrument’s relevance. The involvement of international experts from various institutions globally providing feedback related to the content, contributes to the instrument’s credibility.

However, the study also has limitations, homogeneity of the participants as they are mainly from western cultures and educated to bachelor level affects the generalisability of the findings. The limited sample size represents a limitation, as it may have constrained the accurate estimation of the effect on empowerment in primiparous and multiparous women. In line with this, relatively few participants completed the MPower scale before giving birth to their first child, which could also lead to underestimation of the correlation between parity and empowerment.

### Recommendations

Future research should validate the MPower instrument in larger and more diverse populations, including women from low- and middle-income countries, culturally diverse groups, and nulliparous women. This would strengthen its generalisability and allow clearer comparison across demographic subgroups. Further cross-cultural testing is also needed to determine whether the construct of empowerment is universally applicable or requires contextual adaptation.

Refinement of the instrument should focus on simplifying language and reducing item burden which would improve feasibility and accessibility in both research and clinical settings. Testing discriminant validity against related constructs such as self-efficacy and confidence is also recommended. Further additional work, using varied study designs is required to explore factors that influence empowerment and to investigate possible causal pathways.

Finally, given the consistent association between midwifery continuity of care and higher empowerment, research should examine the MPower’s utility for monitoring empowerment in clinical practice and informing policy initiatives.

## Conclusion

The development and validation of the MPower instrument represent a significant advancement in the actualisation of evidencing perinatal empowerment, reliable for use in high income settings. This instrument allows for a comprehensive assessment of internal and external dimensions that contribute to women’s empowerment during the perinatal period. The MPower scale’s robust psychometric properties make it a trustworthy instrument for healthcare providers, and researchers in high income countries. However, further validation is required, to comprehensively assess the instrument’s dimensional structure and overall validity in other populations.

## Data Availability

The data underlying the results presented in the study are available from at Edith Cowan University

